# Level of Knowledge in the COVID-19 Pandemic: A Cross-Sectional Survey of Canadian Medical Students

**DOI:** 10.1101/2020.10.07.20208801

**Authors:** Kacper Niburski, Rachel Vaughan, Elitsa Papazova, Keith J. Todd

## Abstract

**Background:** During health crises medical education is often derailed as was the case during the current COVID-19 pandemic. Medical trainees face the daunting task of having to gather, filter and synthesize new information about the evolving situation often without the standardized resources they are used to.

**Methods:** We surveyed Canadian medical students, in the hardest hit province of Quebec, on how they were acquiring knowledge as well as what they knew of the pandemic. Google Forms was used, with the survey being distributed to each medical school in Quebec (McGill, ULaval, Udem) both through email and through social media pages for each class year. Two analyses, Mann-Whitney and ANOVA tests, were performed for year of study and degree obtained.

**Results:** We received responses from 111 medical students from three universities, which represents 5% of the students invited to complete the survey. Students reported using mass media most frequently (83%) and also had a high rate of use of social media (to gather information about the pandemic. They rated these resources low in terms of their trustworthiness despite the high rates of use (average 2.91 and 2.03 of 5 respectively). Medical students also endorsed using more formal resources like public health information, scientific journals and faculty-provided information that they trusted more, however, they accessed these resources at lower rates. Of note, medical students had correct answered 60% of COVID-19 prevention strategies, 73% clinical correct answers, 90% epidemiological correct answers. Additionally, students who were training in the larger city of Montreal, where the worst of the outbreak was focused, tended to significantly perform better (p<0.0001) than their colleagues who were not located there.

**Conclusion:** These finding indicate a wide use of information resources intended for public consumption rather than more rigorous and trustworthy sources. Furthermore, there seems to be a knowledge gap amongst medical students responding to this survey that suggests an opportunity to improve the delivery of educational content during this rapidly evolving pandemic.

## Introduction

Medical students serve as an important part of the healthcare system’s future. During pandemics, however, their role is largely unclear. This is evident in Quebec medical schools, where during the SaRS-COV 2 (COVID-19) pandemic the third-year medical students were removed from clinical duties; where second-year medical students were exempt from their transition to clerkship practices; and where fourth-year students were not given any clear role in participating in healthcare services during this evolving crisis (1).

Previous studies have noted that healthcare practitioners including students would be willing to volunteer in the event of a pandemic, a number which increased if given protective garments (2, 3). Many attribute this willingness to a sense of professional obligation, as well as a need to be present when patients are most vulnerable even if students often feel their knowledge is lacking (4, 5, 6).

In COVID-19 there has been confusion regarding the type of work available to students, their clinical duties, and the best means to assist, Partly, this is due to the evolving nature of the pandemic, with ever-changing guidelines and recommendations (7, 8, 9). A knowledge gap has been noted in other countries amongst medical students with regards to COVID-19 (10, 11, 12). Moreover, in previous pandemics, it was medical students that showed the greatest difference in knowledge (13, 14).

Due to this gap in knowledge in other countries and decentralization of the knowledge that does exist, we were curious to investigate the perceptions and knowledge of medical students in Quebec, Canada. The province of Quebec held the distinction of having the highest prevalence rate for COVID-19 infections of all the provinces in Canada. The primary objective of this study is, therefore, to assess the level of COVID-19 clinical knowledge among medical students. This includes determining if students understand the lab findings, clinical presentation, and if they believe the myths in COVID-19. The secondary objectives include understanding what information sources students turn to, and the perceived trustworthiness of those sources. Finally, we were curious as to whether there is an association between students’ knowledge and prior degrees obtained.

## Methods

### Survey Development

The survey was modified off of a validated survey used during the H1N1 pandemic, by University of Alberta (13) (Appendix A, English Version). This previous survey concentrated on the H1N1, however. We included more COVID-19 related questions such as epidemiology, prevention, and clinical information, as well as questions that assessed perceived trust in informational sources. We further asked questions about the students’ backgrounds, questions about laboratory findings, symptoms, treatment, and others about myths and early conspiracy theories. The questions themselves were verified to be the most up-to-date, using multiple sources and public health agencies to obtain accurate information including figures of COVID-19 prevalence (15, 16, 17).

The survey was created on Google Forms (Google Form, Mountain View, California) with 55 questions (Appendix A). 5-point Likert scales, with 5 being “always true”, 4 being “somewhat true”, 3 being “neutral”, 2 being “somewhat false”, and 1 being “always false” were used. This scale for some questions is necessary given the evolving nature of COVID-19 (15, 16). For example, whether COVID-19 can be transferred via infected animals was considered an evolving question with responses true, somewhat true, and neutral being valid as minks and humans have been shown to have some cross-infective potential with unclear consequences (23).

True and false questions were available, as was the choice, “We don’t know.” The survey was internally tested against four medical students, and one staff physician. The survey was available in English and French. Only one response per student was allowed to avoid duplicate or contradictory information. During the ethics review process, it was requested that we remove a question on the trustworthiness of faculty documents, therefore, students do not rate their trust of faculty documents but can indicate it as a source of information.

Ethics approval was received (A06-B41-20A) from McGill University.

### Data Collection

The survey was distributed to each medical school in Quebec both through email and through social media pages for each class year (i.e. Med 1s, Med 2s, etc). Each school distributed the survey once, either in email or social media. Links were directly available in the associated post.

The survey was sent out at varying times by the individual schools. McGill sent out the survey to its students on July 2nd, Université Laval on July 7th, and Université de Montréal on July 8th. Université de Sherbrooke did not respond to any requests, despite being contacted numerous times. The survey was closed on August 20th, 2020 for all schools.

### Statistical Analysis

Only completed responses were collected for data analysis. All data was funneled into Excel (Microsoft, Richmond, Virginia). Mean scores were calculated. Independent sample t-tests were performed for differences in prevalence of COVID-19 (Montreal, with schools Université de Montréal and McGill University, vs Quebec City, with Université Laval). Two analyses, Mann-Whitney and ANOVA tests, were performed for year of study and degree obtained. For the multivariate testing, Likert responses always true and somewhat true were one group, and responses either true or false, somewhat false, and always false were combined according to a method by Bo Gong (24). STATA (StataCorp, Release V.15.1, College Station, Texas, USA) was used for all statistical analyses. P <0.05 were considered significant.

The denominator for response rates was calculated using known class sizes (18), with an observed and expected dropout rate of students at approximately 5% (19). This consideration incorporates those who would see the survey on an online medium. A conventional response rate is usually calculated on national enrolment, estimated to be about 2.8% (20, 21).

## Results

### General

Three of the four Quebec medical schools were represented by responses to the survey. One hundred and eleven students participated from a total of 2148 (5%), which is higher than national averages for one-time surveys (20, 21). Table 1 details the characteristics of the participants. Forty-eight (43.2%) were from McGill University, 34 (30.6%) from Université de Montréal (UdeM), and 29 (26.1%) from Université Laval (ULaval).

**Table 1.**
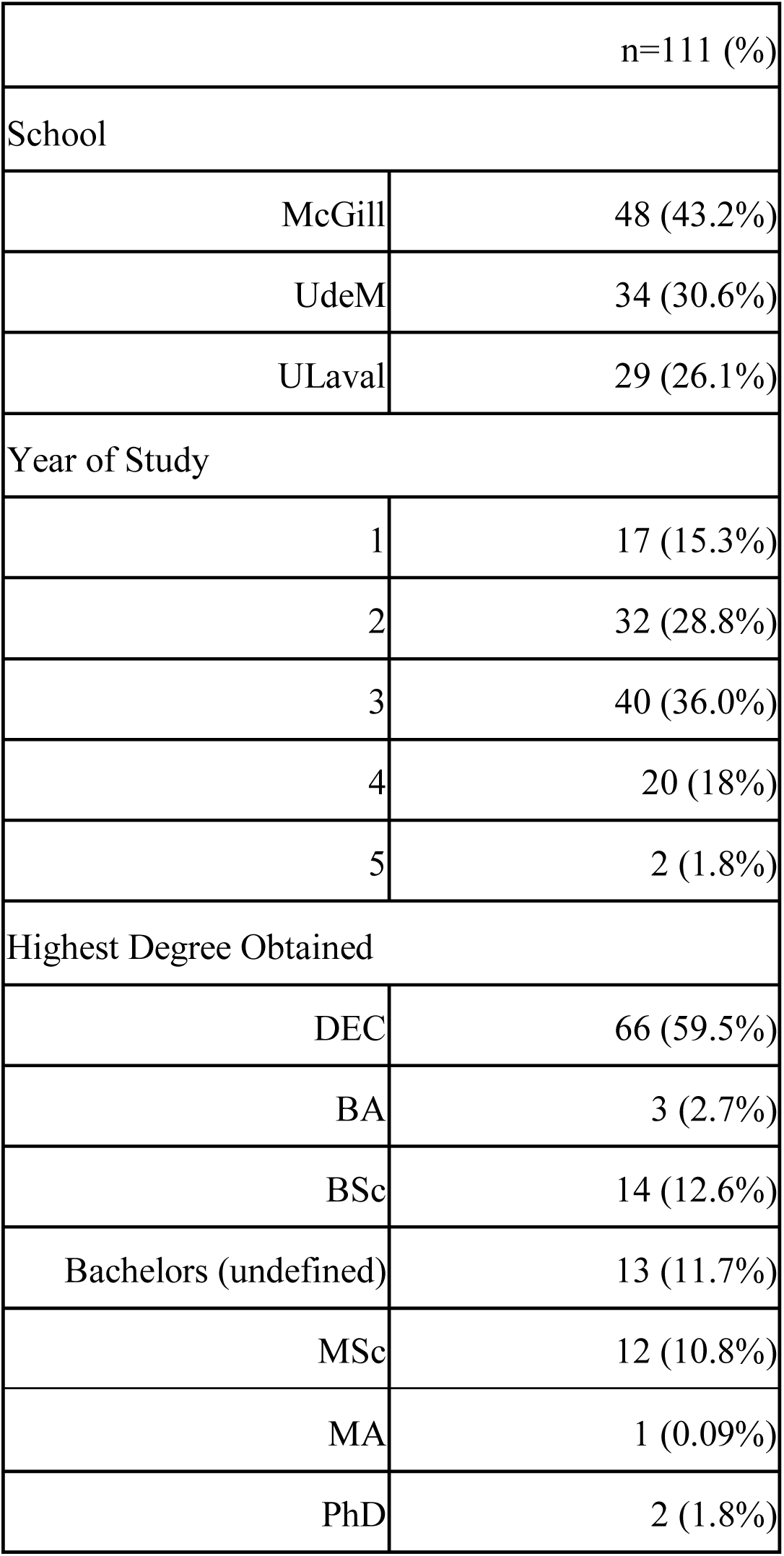
General characteristics of study participants (n=111)

The largest group were from year 3 (40, 36%), followed by year 2 (32, 28.8%). Sixty-six students (59.5%) had a Quebec Diploma of College Studies (DEC), which is given to a student after completion of a college program (22). Thirty had some form of bachelor’s degree, with 14 (12.6%) having a Bachelor of Science and 13 (11.7%) an undefined bachelor’s degree. Only 15 had an advanced graduate degree, with 12 (10.8%) having a Master of Science.

### Source and Perceptions of Information

Medical students look to a number of different resources for information (Fig. 1). Responses were grouped into sources widely available to the public (Fig 1a, c) and those that could be considered for use by health professionals (Fig 1b, d). The most widely used public resource was the mass media (average [AVG] 4.07, standard deviation [SD] 0.83), followed by social media (AVG 3.09, SD 1.33). Family members were not a widely utilized source of information (AVG 2.80, SD 1.21) (Fig. 1a). The most widely accessed scientific resource were public health agencies (AVG 3.94, SD 1.05), followed by scientific journals (AVG 3.47, SD 1.14), and finally faculty documents (AVG 2.59, SD 1.30) (Fig. 1b).

**Figure 1a.**
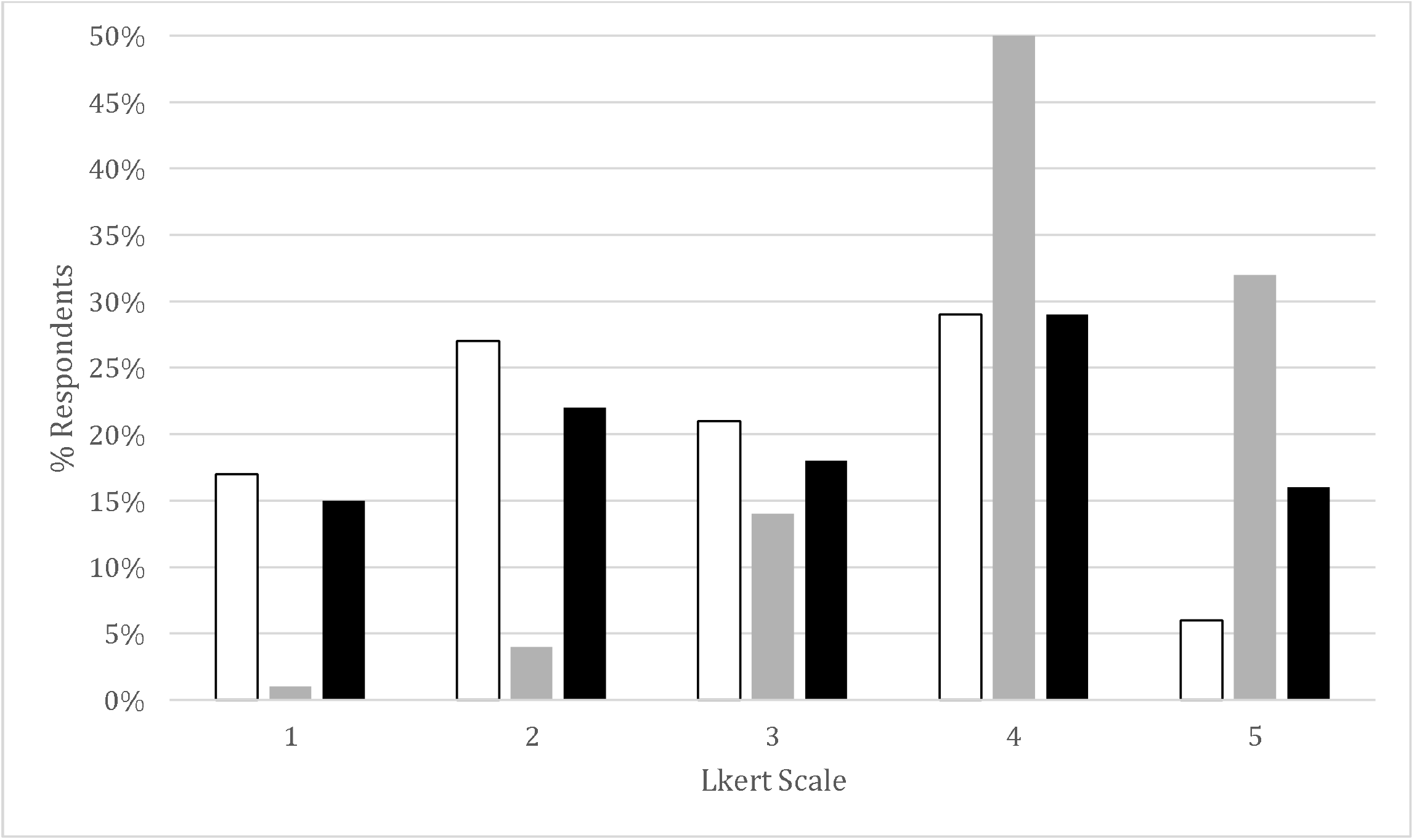
Sources of information for mass consumption, where students have reported learning about COVID-19. Likert scores of 1 indicate being untrue while Likert scores of 5 indicate very true. Family members are white bars, mass media are grey and social media, black.

**Figure 1b.**
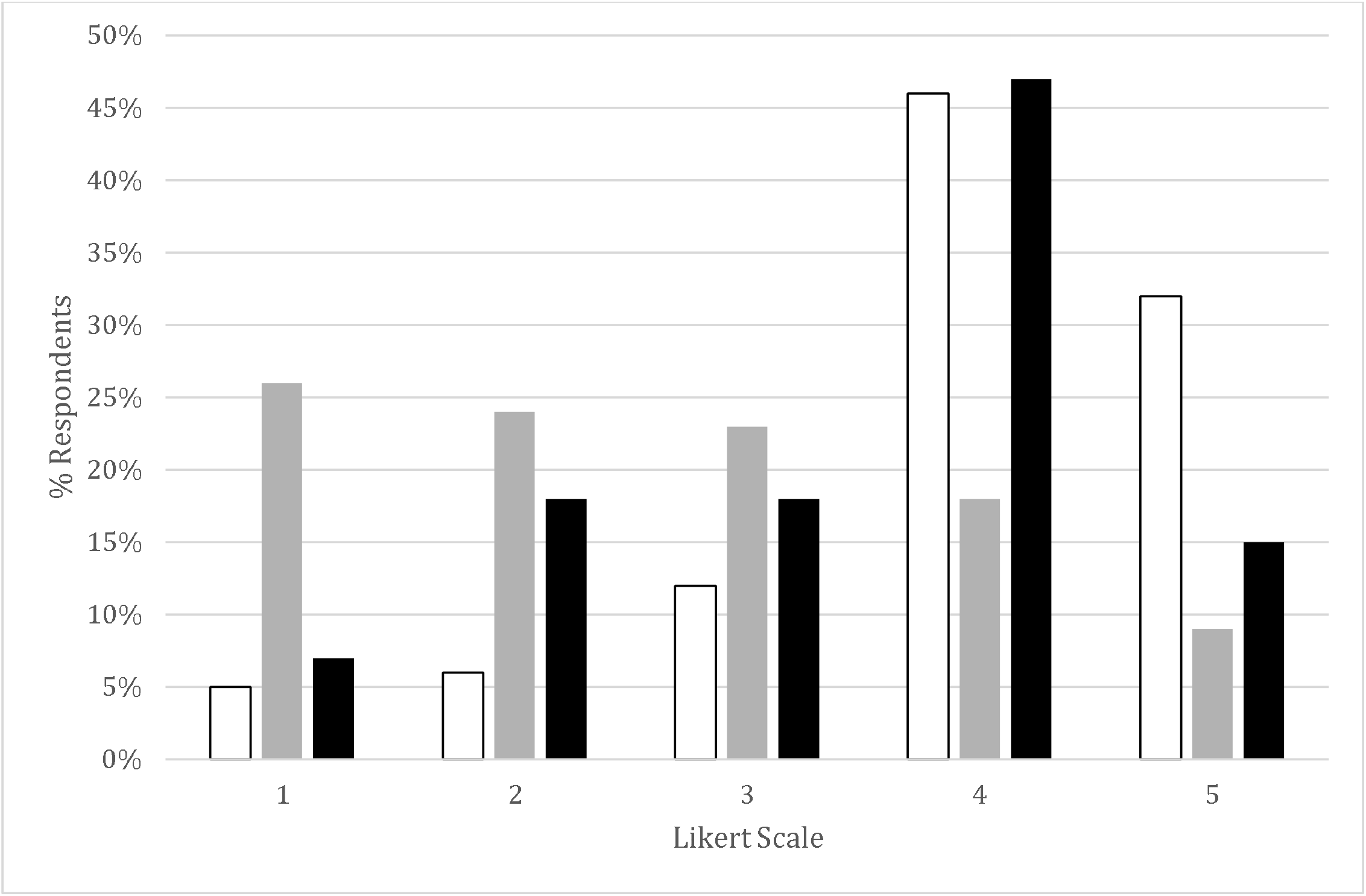
Sources of information directed towards health professionals where students have reported learning about COVID-19. Public health are white bars, faculty documents are grey, and scientific journals, black.

**Figure 1c.**
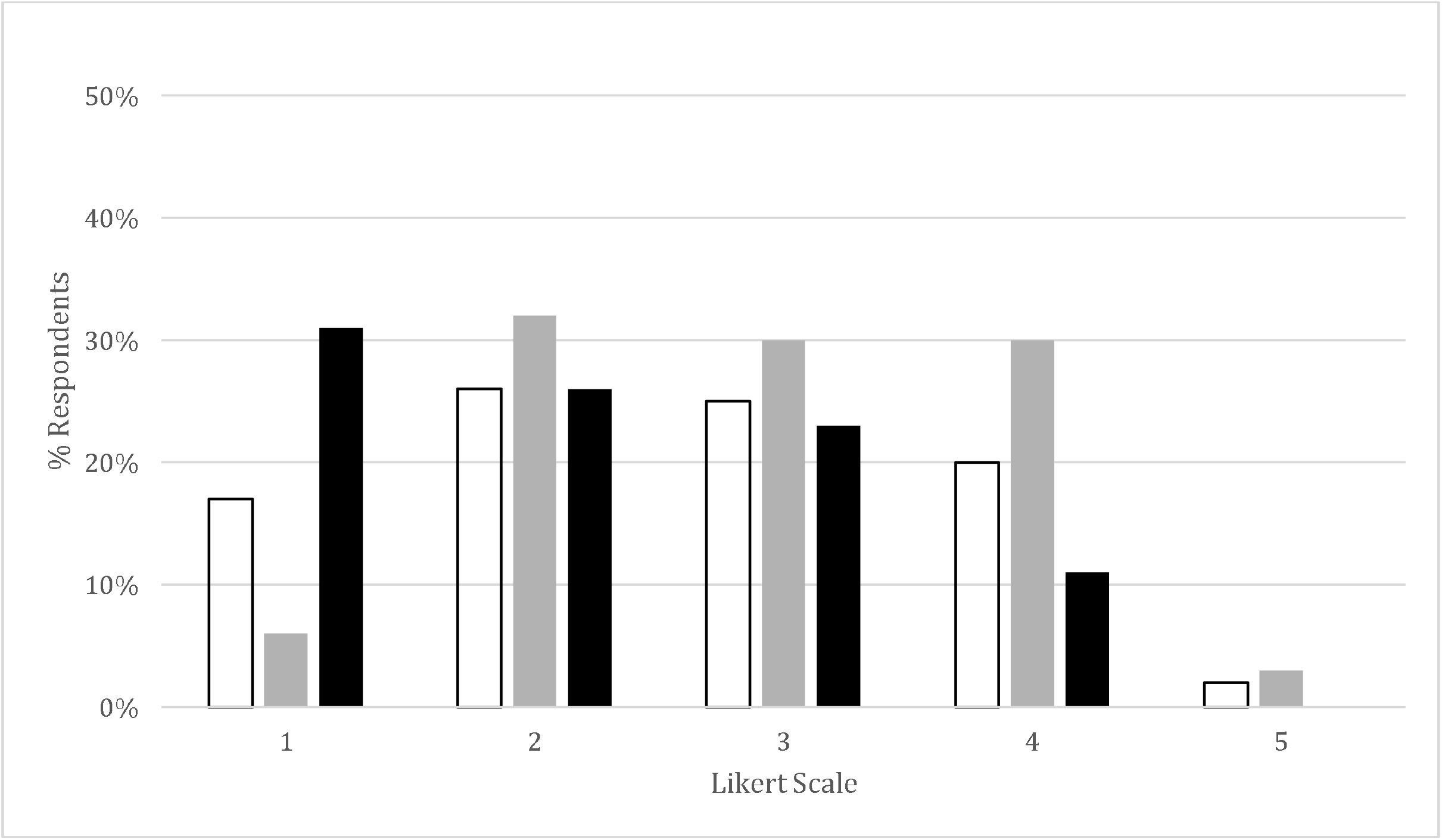
Trust in publicly available information where students have reported learning about COVID-19. Family members are white boxes, mass media are grey, and social media, black.

**Figure 1d.**
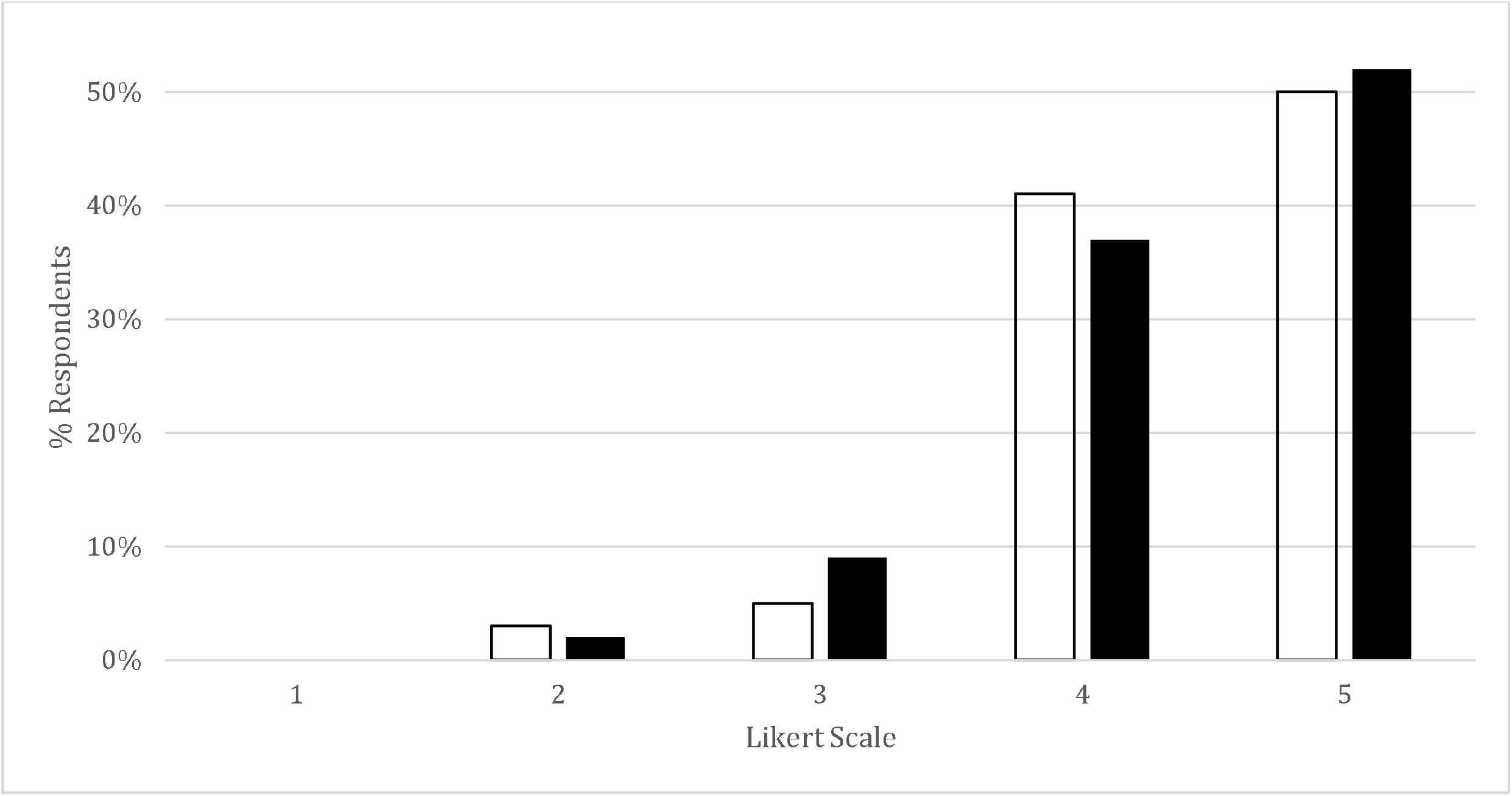
Trust in official information where students have reported learning about COVID-19. Public health are white boxes, and scientific journals, black.

Trustworthiness of publicly available resources were generally rated as not being trustworthy (Fig. 1c). Mass media come closest to a neutral rating (AVG 2.91, SD 0.99). The most trustworthy sources of information were scientific journals (AVG 4.40, SD 0.72) and public health agencies (AVG 4.39, SD 0.71) (Fig 1d). We also compared the use of a given source vs the reported level of trust (Fig. 2). Notably, mass media is most used but not widely trusted whereas public health documents were accessed at similar rates but were highly trusted.

**Figure 2.**
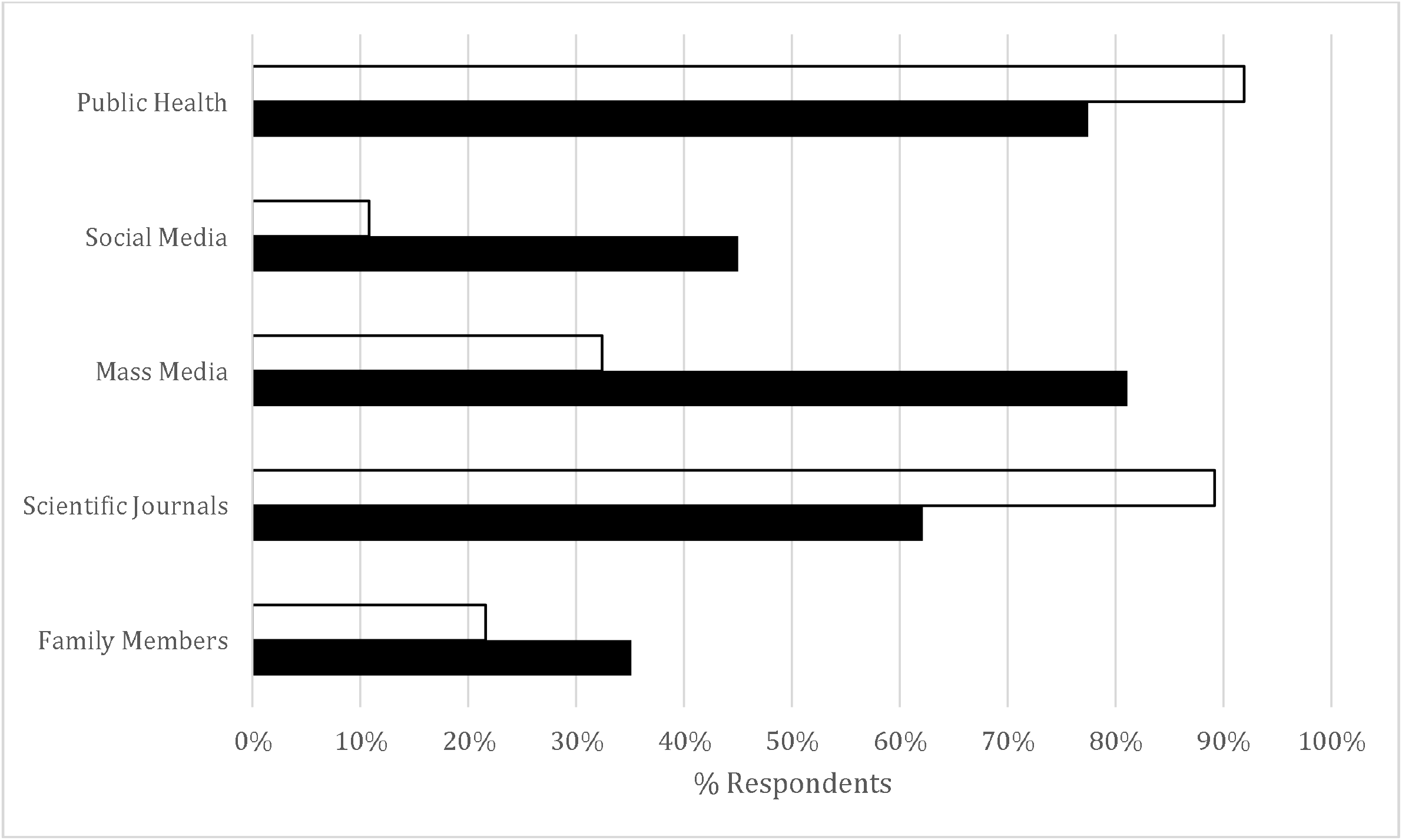
Informational sources vs their associated trust. White bars illustrate the % of participants who trust (greater than a neutral value on the Likert-scale) and black bars indicate the % of participants who use the informational source (greater than a neutral value on the Likert-scale).

### Transmission, Prevention, and Treatment Knowledge

Students were asked to respond to questions about the transmission, prevention, treatment and basic epidemiology of the pandemic. Question categories were grouped and reported as the percentage of correct answers (Table 2). Student respondents did well with 77% correct answers on average for questions about transmission. They scored 69% on questions of prevention and only 64% when it came to questions related to treatment.

**Table 2.**
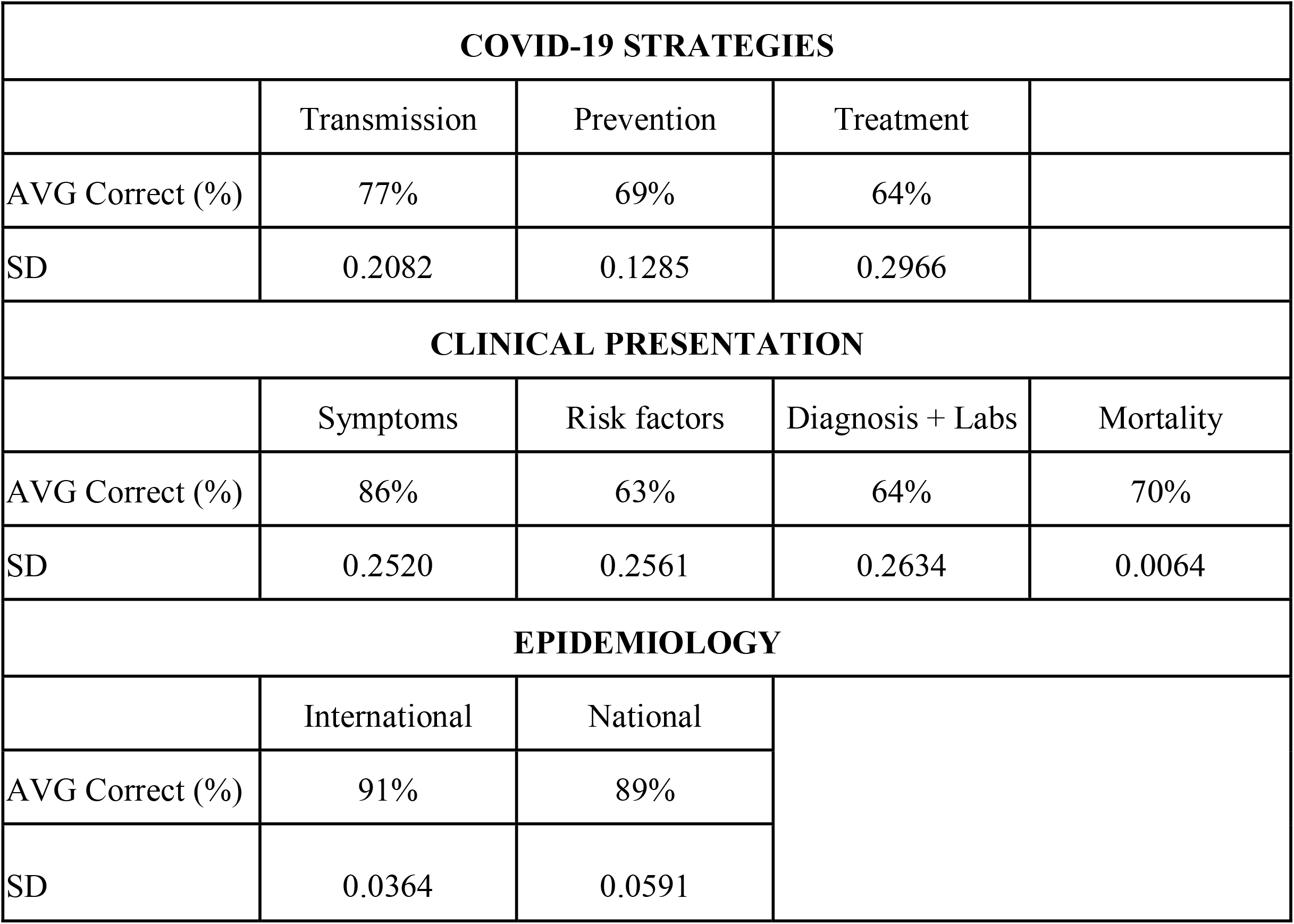
Responses to questions related to treatment, prevention, transmission, clinical information, diagnosis, and epidemiology at an international and national level were tabulated and presented as average +/- standard deviation of correct responses.

Some specific questions about prevention revealed interesting results. For instance, 72 students (64.9%) knew that ten seconds of handwashing was not recommended, while 100 (90.1%) students responded correctly that 20-seconds was recommended. Other prevention strategies were also relatively well known: 88 students (79.3%) stated that quarantine was effective, 72 (64.9%) stated masks were effective, however, only 63 (56.8%) knew that gloves were not necessarily effective.

Treatment-related questions varied in accuracy of response and generally students did more poorly on these questions. For example, currently approved treatments such as dexamethasone (17, 15.3%) and remdesivir (23, 20.7%) were not widely known to be approved. However, unverified medical treatments such as azithromycin (92, 82.9%) and chloroquine (69, 62.2%) were known to be ineffective. Of note, medications like chloroquine, which have been noted to have an uptick in media attention, received more “we don’t know” (34, 30.6%) and statements of efficacy (8, 7.2%) than other medications.

### Clinical Understanding

Questions related to clinic presentation, diagnostics and mortality were grouped. Correct answers were tabulated and are expressed as average correct +/- standard deviation (Table 2). Questions about clinical symptoms were answered with an overall 86% correct. Interestingly, a question about the most common symptom, cough, fever or anosmia was split with 46 respondents indicating fever (41.4%) and another 42 (37.8%) indicating cough as the most common symptom. The majority of students knew that COVID-19 patients could be asymptomatic (107, 96.4%). Generally, questions related to risk factors and diagnosis were more poorly done with average correct rates of 63% and 64% respectively. In terms of mortality rates, most knew that influenza has a lower death rate (77, 69.3%), and Ebola was higher (78, 70.2%).

### Epidemiology

Questions related to international and Canadian epidemiology were grouped. Average correct responses were calculated demonstrating good basic understanding of international and national epidemiology with 91% and 89% correct respectively (Table 2). Interestingly, ten students (9.0%), stated that Ontario had the highest prevalence of COVID-19 despite residing in the hardest hit province of Quebec.

### Comparison Groups

#### Prevalence of COVID-19 in the Community

Responses were divided according to where individuals attended medical school. Respondents who attended medical school in Montreal (UdeM or McGill) were grouped and compared those from ULaval in Quebec City. Overall, responses were largely similar. There were a few cases where statistically significant differences were recorded. In these cases responses from students attending medical school in Montreal tended to be more accurate (Table 3). ULaval students showed more incorrect assumptions regarding COVID, including that COVID-19 can be prevented by current vaccinations (AVG 2.172 vs 1.488, p=0.0439), prevented by natural vitamins (AVG 1.862 vs 1.317, p=0.0092), and prevented by drinking alcohol (AVG 1.448 vs 1.158, p=0.0399). As a whole, however, the answers were still largely on the correct side of the Likert-scale.

**Table 3.**
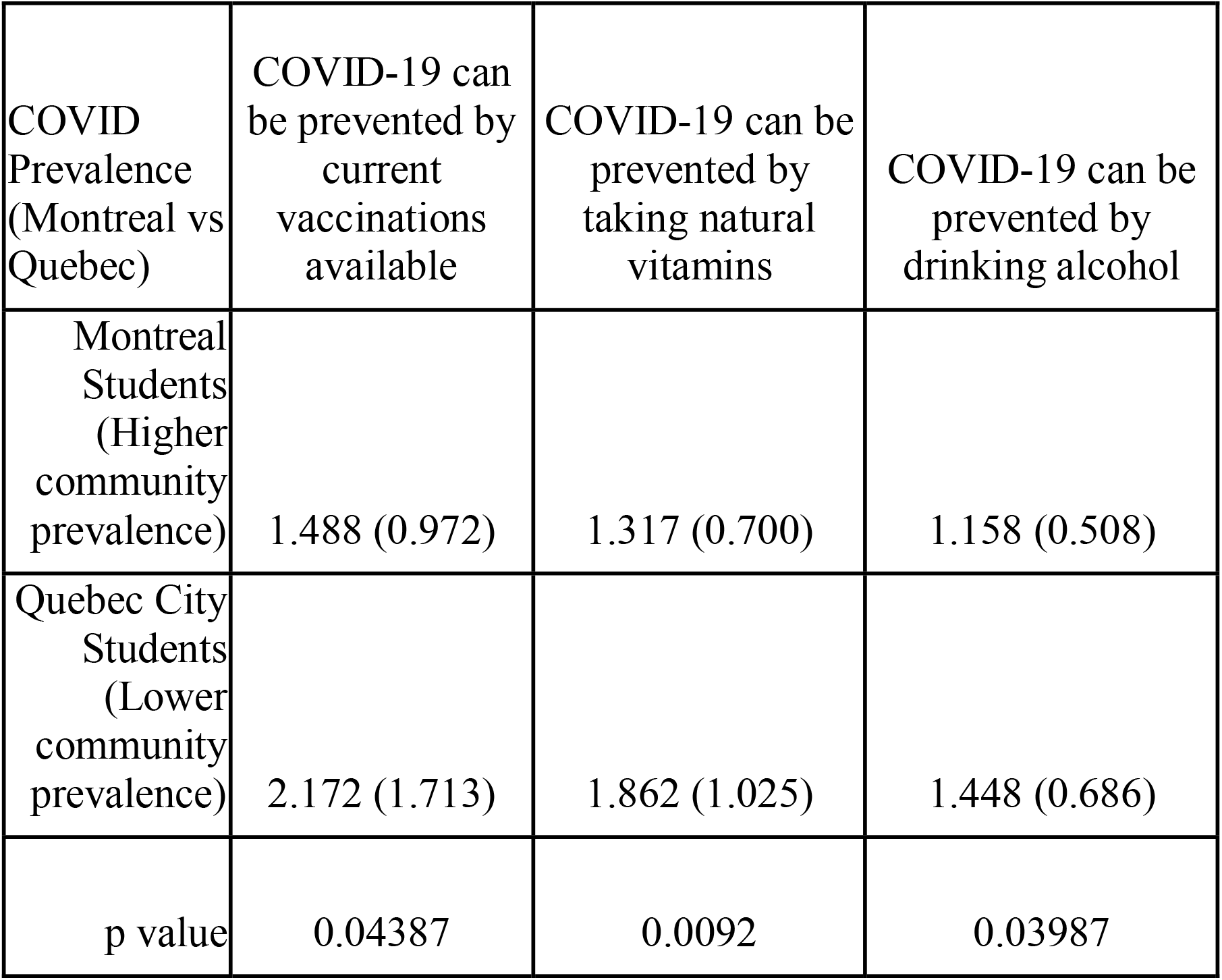
Responses divided by location of medical school (with-in Montreal vs Quebec City). Results provided in average (standard deviation). p value given significant above 95%.

#### Year of Study

Comparing by year of study yielded no significant difference in most knowledge, except for the fact whether chloroquine can prevent COVID-19 (p=0.0043). Third year students scored best (AVG 1.625, SD 0.867), followed by fourth year students (AVG 1.70, SD 0.922). Second year students scored the worst (AVG 2.438, SD 1.014). Furthermore, as the year of study progressed, students were less likely to learn from their family members (p=0.0029) or think of their families as trusted sources (p=0.0004) (Table 4).

**Table 4.** Responses by year of study presented as average +/- standard deviation for questions showing significant differences (p<0.05). Results provided in average (standard deviation).

| Year of Study | I have been learning about the pandemic from family members | Family members are a trusted source for pandemic information. | COVID-19 can be prevented by antivirals such as chloroquine |
| --- | --- | --- | --- |
| 1 | 3.359 (1.412) | 3.058 (1.088) | 2.352 (1.32) |
| 2 | 3.188 (1.148) | 2.906 (1.027) | 2.438 (1.014) |
| 3 | 2.425 (1.083) | 2.4 (1.008) | 1.625 (0.867) |
| 4 | 2.35 (1.039) | 1.8 (0.696) | 1.7 (0.922) |
| 5 | 4.0 (0) | 2.0 (0) | 2.5 (0.707) |
| p value | 0.0029 | 0.0004 | 0.0043 |

#### Degree Specific

We further compared individuals who completed university degrees (Bachelors, Master’s, and PhD’s) vs those without university degrees (DEC) (Table 5). DEC holders showed significantly more trust (AVG 2.59 vs 2.44) in family, less use of scientific journals (AVG 3.17 vs 3.93), and less trust in public health (AVG 4.26 vs 4.6). Students entering medical school with a DEC also responded less accurately, on average, to questions on treatments such as the use of dexamethasone (AVG 1.89 vs 2.2) and remdesivir (AVG 2.15 vs 2.44).

**Table 5.** Responses to questions relating to transmission separated by the level of education obtained prior to entering medical school, presented as average +/- standard deviation.

| **SOURCE OF INFORMATION** |  |  |  |  |  |
| --- | --- | --- | --- | --- | --- |
|  | Family | Scientific Journals | Social Media | Public Health | Total |
| DEC | 2.88 (1.16) | 3.17 (1.20) | 3.20 (1.34) | 3.83 (1.10) | 3.27 |
| Higher Education | 2.68 (1.29) | 3.93 (0.89) | 2.93 (1.32) | 4.09 (0.95) | 3.41 |
| p value | 0.00001 | 0.00001 | 0.00001 | 0.00001 |  |
| **TRUST OF INFORMATION** |  |  |  |  |  |
|  | Family | Scientific Journals | Public Health |  | Total |
| DEC | 2.59 (1.08) | 4.26 (0.81) | 4.26 (0.81) |  | 3.70 |
| Higher Education | 2.44 (1.01) | 4.6 (0.54) | 4.6 (0.50) |  | 3.88 |
| p value | 0.0293 | 0.00229 | 0.00169 |  |  |
| **PREVENTION** |  |  |  |  |  |
|  | Handwashing 10 seconds | Quarantine effective | Mask in public | Gloves use | Total |
| DEC | 2.38 (1.26) | 3.82 (1.09) | 3.44 (1.07) | 2.30 (1.11) | 2.99 |
| Higher Education | 1.89 (0.98) | 4.44 (0.89) | 4.04 (0.90) | 2.58 (1.25) | 3.24 |
| p value | 0.00001 | 0.00001 | 0.00001 | 0.00001 |  |
| **TREATMENT** |  |  |  |  |  |
|  | Azithromycin | Dexamethasone | Remdesivir | Staying hydrated | Total |
| DEC | 1.62 (0.93) | 1.89 (1.12) | 2.15 (1.26) | 1.86 (1.12) | 1.42 |
| Higher Education | 1.33 (0.67) | 2.2 (1.27) | 2.44 (1.34) | 1.71 (1.18) | 1.92 |
| p value | 0.00001 | 0.00001 | 0.00001 | 0.00001 |  |

## Discussion

This is the first study, to our knowledge, that looks at the informational sources used by medical students in Canada to acquire new knowledge during the COVID-19 pandemic. The survey results suggest that medical students obtain much of their information with social media and mass media, noting, however, that both are not trustworthy. Scientific journals and public health institutions were the most trustworthy, but not as widely accessed for information. One consideration for this result may be due to the higher levels of trust for the media amongst Canadians when compared to the United States (25). It is, of course, important to consider the ease of access for the information, which no doubt, contributes to high levels of social and mass media use.

Previous international research has noted that among residents and students, there is a gap in knowledge surrounding COVID-19 (10, 11, 12), though in Canada, this is the first such investigation. It is certainly not surprising that there were some knowledge gaps as best practices were evolving quickly during the course of the study. Furthermore, the students surveyed were not allowed to be actively involved in the care of patients. A key question for educators is how to help students stay abreast of current knowledge during a rapidly evolving crisis.

Still, the students did well but there are notable gaps in knowledge of treatment and prevention. Without a comprehensive understanding of pandemics, and with widespread use of unverified sources of information, there will be ongoing difficulty in providing accurate, evidence-based care by medical students (35). Some student-led initiatives, such as the Harvard COVID curriculum (26, 27) or a COVID-19 tracker (28), offer some sources for knowledge; however, they are not widespread.

The apparent knowledge gap amongst students certainly does bring to question whether there was a missed educational opportunity. Would students have been more engaged in learning if they were actively involved in care? The stoppage of training that medical students in Canada experienced due to the pandemic could have possibly been turned into a global health and pandemic elective, thereby encouraging learning through in-person, workplace learning (36).

Furthermore, it is important to note that previous studies have shown that social media offer opportunities for innovation in medical education (29). Often, there are associations with improved knowledge (30), and greater collaboration is possible (31). While many of the stories of COVID-19 and social media exacerbates myths and conspiracy (32), others in medical education used social media to create virtual committees and hubs of knowledge (33). Further research could look at the effect of the institution of Quebec-specific COVID-19 curriculum and its effect on knowledge, as well as the role of social media in COVID-19 among medical students.

There are certainly limitations to this study. The data is self-reported, and subject to recall bias by the students. It is also a relatively small number of students from three schools all in the province of Quebec. The generalizability of the results are unclear. Despite this, this survey does provide interesting insight into knowledge and knowledge acquisition by medical students during this unprecedented time. Additionally, though the lockdown measures were mandated by law to be the same provincially in Quebec, their local application may have differed (34). Such a difference, even small, may help explain the differences in Quebec City and Montreal students due to more exposure to the news or other sources by Montreal students compared to lower-prevalence students in other geographical locations.

## Conclusion

As the first study of its kind in Canada, the results suggest there are knowledge gaps and wide-spread use of unverified and poor informational sources like social media amongst medical students in Quebec, Canada. Temporary interruptions to the normal medical curriculum and global health crises provide different learning opportunities for health professionals. There is perhaps an un-tapped opportunity for medical schools to plan for future disruptions and how best medical students can be engaged and their learning enhanced.

## Supporting information

Appendix A, Survey

## Data Availability

The datasets used and/or analysed during the current study are available from the corresponding author on reasonable request.

## Declarations

Ethics approval was received (A06-B41-20A) by McGill University, with no more than minimal risk and in accordance with Articles 2.9 and 6.12 of the 2nd Edition of the Canadian Tri-Council Policy Statement of Ethical Conduct for Research Involving Humans (TCPS 2 2018) and U.S Title 45 CFR 46, Section 110 (b) paragraph 1. All protocols are carried out in accordance with relevant guidelines and regulations and a statement confirming that the informed consent was obtained from the all participants for the study

### Consent for publication

Not applicable as participants were anonymous.

### Authors contributions

Kacper Niburski: designed, performed data analysis, interpreted data and writing the manuscript.

Rachel Vaughan: helped with design and writing the article

Elitsa Papazova: helped with design and writing the article

Keith Todd: Helped with design, analysis, interpretation and writing the manuscript

## Acknowledgement

Not applicable.

