## Appendix A, Survey for "Level of Knowledge in the COVID-19 Pandemic: A Cross-Sectional Survey of Canadian Medical Students"

| 1 | What year of study are you? |
| --- | --- |
| 2 | What is your highest degree obtained? |
| 3 | On a scale from 1 to 5, 5 being always true, 3 being unknown or neutral, and 1 being always false, I have been learning about the pandemic from family members. |
| 4 | I have been learning about the pandemic from scientific journals. |
| 5 | I have been learning about the pandemic from mass media. |
| 6 | I have been learning about the pandemic from social media. |
| 7 | I have been learning about the pandemic from documents released by the faculty of medicine. |
| 8 | I have been learning about the pandemic from local, provincial, and federal public health agencies. |
| 9 | I have been avoiding learning about the subject. |
| 10 | The media is a trusted source for pandemic information. |
| 11 | Social media is a trusted source for pandemic information. |
| 12 | Family members are a trusted source for pandemic information. |
| 13 | Scientific journals are a trusted source for pandemic information. |
| 14 | Public health agencies are a trusted source for pandemic information. |
| 15 | On a scale from 1 to 5, 5 being always true, 3 being unknown or neutral, and 1 being always false, transmission of COVID-19 can occur through close contact with infected persons? |
| 16 | Transmission of COVID-19 can occur through blood transfusion? |
| 17 | Transmission of COVID-19 can occur through cough/sneeze? |
| 18 | Transmission of COVID-19 can occur through touching surfaces? |
| 19 | Transmission of COVID-19 can occur through contact with infected animals? |
| 20 | COVID-19 can be significantly reduced with hand washing for ten seconds? |
| 21 | COVID-19 can be significantly reduced with hand washing for twenty seconds? |
| 22 | COVID-19 can be significantly reduced by coughing/sneezing into elbow? |
| 23 | COVID-19 can be prevented by current vaccinations available? |
| 24 | COVID-19 can be prevented by antibiotics such as azithromycin? |
| 25 | COVID-19 can be prevented by antivirals such as chloroquine? |
| 26 | COVID-19 can be significantly reduced by quarantine? |
| 27 | COVID-19 can be significantly reduced by wearing a mask in public? |
| 28 | COVID-19 can be significantly reduced by wearing gloves in public? |
| 29 | COVID-19 can be prevented by taking natural vitamins? |
| 30 | COVID-19 can be prevented by taking dexamethasone? |
| 31 | COVID-19 can be prevented by taking remdesivir? |
| 32 | COVID-19 can be prevented by staying hydrated? |
| 33 | COVID-19 can be prevented by drinking alcohol? |
| 34 | Choose the best answer: the most common symptom of a symptomatic COVID-19 infection? |
| 35 | All patients show symptoms of a COVID-19 infection? |
| 36 | Influenza has a higher higher mortality rate than corona virus? |
| 37 | Ebola has a higher higher mortality rate than corona virus? |
| 38 | Males are more at risk of infection for COVID-19? |
| 39 | Radiographic data is necessary for diagnosis of COVID-19? |
| 40 | Lab findings include lymphopenia for COVID-19 infections? |
| 41 | Lab findings include elevated d-dimer for COVID-19 infections? |
| 42 | Nasopharyngeal swabs and results can be processed in an hour for COVID-19? |
| 43 | Steroids are useful for treatment of COVID-19? |
| 44 | All patients will require ICU intervention for COVID-19? |
| 45 | All patients will require ICU intervention for COVID-19? |
| 46 | The majority of COVID-19 patients will require ventilator support? |
| 47 | The most at risk are the middle aged for COVID-19? |
| 48 | The most common comorbidity of a COVID-19 infection is rheumatological? |
| 49 | If you cannot hold your breath for ten seconds, you have COVID-19? |
| 50 | The source of the global COVID-19 infection was America? |
| 51 | The source of the global COVID-19 infection was China? |
| 52 | Canada has the lowest COVID-19 infection rate of all countries? |
| 53 | Ontario is the province with the most COVID-19 cases? |
| 54 | British Columbia is the province with the most COVID-19 cases? |
| 55 | Quebec is the province with the most COVID-19 cases? |
